# Opportunistic CT Attenuation Biomarkers of Anemia Are Associated With Impaired Myocardial Flow Reserve and Cardiovascular Outcomes

**DOI:** 10.64898/2026.05.14.26353239

**Authors:** Robert J.H. Miller, Aakash Shanbhag, Jirong Yi, Jacek Kwiecinski, Paul Kavanagh, Giselle Ramirez, Mark Lemley, Assiata Kamagate, Leandro Slipczuk, Mark I. Travin, Erick Alexanderson, Isabel Carvajal-Juarez, René R.S. Packard, Mouaz Al-Mallah, Andrew J. Einstein, Wanda Acampa, Stacey Knight, Viet T Le, Steve Mason, Samuel Wopperer, Panithaya Chareonthaitawee, Thomas L. Rosamond, Robert A DeKemp, Ronny R Buechel, Daniel S. Berman, Damini Dey, Marcelo F. Di Carli, Piotr J. Slomka

**Author notes:** Joint first authors (Robert J.H. Miller, Aakash Shanbhag). **Address for Correspondence**: Piotr Slomka, PhD, Cedars-Sinai Medical Center, 6500 Wilshire Boulevard, Los Angeles, California 90048.

## Abstract

**Background:** Anemia is an established marker of cardiovascular disease severity and risk which leads to elevations in resting myocardial blood flow (MBF) and impaired myocardial flow reserve (MFR) in patients without obstructive coronary artery disease (CAD). Anemia can potentially be detected opportunistically from blood pool density changes on computed tomography (CT) imaging.

**Objectives:** We evaluated relationships between chamber density measurements with hemoglobin, positron emission tomography (PET) findings, and cardiovascular events.

**Methods:** We included 33460 patients from 13 sites in the REFINE-PET who underwent PET and 24368 patients undergoing lung cancer screening chest CT. A deep learning model segmented cardiac chambers from CT images, then quantified chamber density. We evaluated the relationship between chamber density measures with resting MBF and MFR, as well as associations with death or myocardial infarction (MI).

**Results:** We included a total of 57,828 patients. A higher density in myocardium compared to left ventricle blood pool was associated with reduced MFR (adjusted odds ratio 3.02 per SD increase, 95% confidence interval[CI] 2.72 – 3.38) and an increased risk of death or MI in (adjusted hazard ratio[HR] 1.38 per SD increase, 95% CI 1.26-1.51). Having myocardial density higher than blood pool density was also associated with cardiovascular death in patients undergoing low-dose chest CT (adjusted HR 1.73, 95% CI 1.20-2.52).

**Conclusions:** In a large multimodality dataset, lower cardiac chamber density is associated with impaired MFR and independently associated with cardiovascular events. These biomarkers can be automatically extracted from CT to provide physiologic insights and potentially guide patient care.

**CONDENSED ABSTRACT:** Anemia is an important marker of cardiovascular disease severity and predictor of cardiovascular events. We utilized deep learning to measure cardiac chamber density on computed tomography imaging as a surrogate for anemia in 57,828 patients from two large multicenter studies. A higher density in myocardium compared to left ventricle blood pool was associated with reduced MFR and an increased risk of death or MI in patients undergoing positron emission tomography, and an increased risk of cardiovascular death in patients undergoing low-dose computed tomography. These biomarkers can provide additionally physiologic insights which could be used to guide patient care.

## INTRODUCTION

Anemia is a critical cardiovascular risk marker, acting both as a measure of cardiovascular disease severity and an independent predictor of cardiovascular events. Anemia is present in up to 60% of patients with heart failure(1), and 10-43% of patients presenting with acute coronary syndrome(2). Anemia is related both to iron deficiency and anemia of chronic disease(3). In turn, the presence of anemia of chronic disease is correlated with inflammation(4), which is a key driver of coronary atherosclerosis progression(5). Anemia is not just a surrogate marker, it is associated with development of heart failure and coronary artery disease (CAD)(6) and an independent predictor of cardiovascular events in healthy cohorts(5).

Myocardial perfusion imaging (MPI) is frequently used to assess patients with known or suspected CAD,(7) but is also influenced by hemoglobin levels. Myocardial blood flow (MBF) measurements are powerful diagnostic and prognostic markers in patients undergoing positron emission tomography (PET) MPI(8). Patients with reduced myocardial flow reserve (MFR) have a two-fold increased risk of cardiovascular death or myocardial infarction (MI). Quantitative myocardial blood flow (MBF) and MFR are increasingly used for diagnosis and risk stratification; however, these measurements are influenced by physiologic factors, including anemia, that are rarely incorporated into image interpretation. In fact, hemoglobin is a major determinant of resting MBF and anemia is associated with an increased probability of having impaired MFR in the setting of preserved stress MBF(9).

Measurement of hemoglobin is not routinely performed prior to MPI. However, previous studies have highlighted the potential for computed tomography (CT) to identify the presence of anemia (10–12). Abbasi et al. demonstrated that visual identification of a differential signal in the aorta (aortic ring sign) or septum (interventricular septum sign) are highly specific for the presence of anemia (87% and 99.2%, respectively)(10). Zhou et al. demonstrated that myocardium density higher than blood pool density is predictive of the presence of anemia in a cohort of 317 patients(12). However, the threshold for mild anemia was only a difference of 4.5 Hounsfield Units (HU), which would be extremely challenging to reliably identify by visual inspection alone(12). Automating quantification of these differences, enabling automated identification of anemia, could potentially help physicians interpret MBF findings and better understand overall risk in patients undergoing PET MPI. Furthermore, this approach could be applied to non-cardiac chest CT imaging as well. Although prior small studies demonstrated that CT attenuation may reflect hemoglobin concentration, it remains unknown whether automated CT-derived biomarkers of anemia are associated with MBF or cardiovascular outcomes in large multicenter populations.

To address this unmet clinical need, we developed and validated a fully-automated approach for segmentation and quantification of chamber density. Using a large, multicenter, international dataset we evaluated associations between chamber density measurements with the presence of anemia, correlations with resting MBF and MFR, as well as independent associations with cardiovascular events in large multicenter population. We also validated the importance of these measures in a separate multicenter cohort of patients undergoing low dose lung CT.

## METHODS

### Patient Populations

A total of 57,828 patients were included in the analysis, with overall study design outlined in the **Central Illustration**. We included 33460 patients from the REgistry of Fast Myocardial Perfusion Imaging with NExt generation PET (REFINE PET) who underwent ^82^Rb and ^13^NH_3_ PET MPI with available CTAC(13). Patients were included from thirteen sites: Brigham and Women’s Hospital (n=7243), Cedars-Sinai Medical Center (n=5992), Columbia University Irving Medical Center (n=1933), Houston Methodist (n=1902), University of Kansas Medical Center (n=603), Montefiore Medical Center (n=1024), Mayo Clinic (n=2010), National Autonomous University of Mexico City (n=381), University of Naples Federico II (n=456), West Los Angeles Veterans Affairs Medical Center (n=1182), Intermountain Healthcare (n=5997), University of Ottawa (n=3981), and Zurich University (n=756). The study was approved by the institutional review boards at each site, with the overall study approved by the Cedars-Sinai Medical Center’s review board.

Additionally, to evaluate the utility of measures in patients undergoing non-cardiac imaging, we included 24368 subjects from the National Lung Screening Trial (NLST; NCT00047385), who were randomized to low-dose chest CT for lung cancer screening(14,15). Only the baseline CT was used in our analysis.

### Clinical Variables

Clinical information was collected at the time of imaging and included: age, sex, body mass index (BMI), family history of CAD, smoking status, history of previous MI, previous revascularization, hypertension, diabetes, and dyslipidemia. Prior CAD was defined as previous MI or revascularization (16). Hemoglobin and hematocrit values within 90 days of PET MPI was available in a subset of patients from two sites. Anemia was defined as hemoglobin <13.2 g/dL in male patients or <11.6 g/dL in female patients and separately as significant anemia if hemoglobin <10.0 g/dL.

### PET Image Acquisition and Quantification

Patients underwent rest and stress PET MPI in accordance with clinical guidelines(17), using either ^82^Rb (n=17294) or ^13^NH_3_ (n=9398) (18). Early dynamic acquisitions were utilized to quantify stress and rest myocardial blood flow (MBF), with myocardial blood flow reserve (MFR) calculated as stress MBF / rest MBF (19). Abnormal MFR was defined as <2 and abnormal resting MBF was considered to be values > 1.2 ml/min/g(19). Stress and rest relative perfusion was quantified using total perfusion deficit (TPD) (20). All PET image quantification was performed at the core laboratory blinded to outcomes with dedicated software (QPET, Cedars-Sinai Medical Center, Los Angeles, California)(20,21). Deep learning coronary artery calcium was quantified as previously described(22,23).

### CTAC Quantification

CTAC scans were acquired according to site-specific PET/CT protocols for all cases as previously described(13). We utilized our previously validated deep learning model to segment cardiac chambers from CTAC images(14,15,24–26). We then quantified median chamber attenuation from the deep learning segmentations. We evaluated left atrium (LA), left ventricle (LV), right atrium (RA), right ventricle (RV), aorta and pulmonary artery (PA) as targets for blood pool measurements and LV myocardium to sample myocardium. Blood pool density < 35 HU was considered abnormal(27). Given the limited size of previous analyses evaluating differences between blood pool and myocardium density, and known overlap between these structures(28), we considered any elevation of myocardium density to blood pool density (difference > 0) to be abnormal.

### Outcomes

The primary outcome was all-cause mortality or myocardial infarction (MI). Death was ascertained from the National Death Index or local administrative databases. Incidence of MI was determined from electronic medical records, with potential events adjudicated by an experienced physician. In the NLST population, we evaluated associations with cardiovascular mortality.

### Statistical Analysis

Continuous variables were summarized as median (interquartile range [IQR]) and compared using a Mann-Whitney U-test. Correlation between chamber density and hemoglobin and hematocrit values were evaluated using Pearson correlation coefficient. Associations with high resting MBF and reduced MFR were assessed using logistic regression. Multivariable models included age, sex, past medical history (hypertension, diabetes, dyslipidemia, family history, smoking, prior CAD), stress TPD, left ventricular ejection fraction (LVEF), and CAC. Univariable and multivariable Cox proportional hazards analyses were conducted to assess associations with the clinical outcomes. Our multivariable model incorporated age, sex, past medical history, stress TPD, LVEF, CAC, and MFR. In the NLST population, associations with cardiovascular mortality were assessed using Fine Gray competing risk analysis. The multivariable model includes age, sex, smoking history and medical history. Analyses were performed using R, version 4.2.3, and Stata version 14.2 (StataCorp, College Station, Texas).

## RESULTS

### Patient Populations

In total, we included 57828 patients in the analysis. We included 33460 patients from REFINE PET with a median age of 67 (IQR 58 – 75) and 15800 (59.2%) male patients **Table 1**. need to say what where the outcomes before going into associations of outcomes and PET Patients who experienced death or MI had higher resting MBF (median 0.98 vs 0.93, p<0.001) and had lower MFR (median 1.88 vs 2.42, p<0.001). We also included 24368 patients from the NLST trial with a median age of 61 and 14455 (59.3%) male patients. outline the outcomes **Table 2**.

**Table 1:** Baseline population characteristics stratified by incidence of death or myocardial infarction (MI). CAC – coronary artery calcium, LVEF – left ventricular ejection fraction, MBF – myocardial blood flow, CAD – coronary artery disease, TPD – total perfusion deficit, MFR – myocardium flow reserve.

|  | **All Patients**<br>**N=33460** | **No Death or MI**<br>**N=25670** | **Death or MI**<br>**N=7790** | **P-value** |
| --- | --- | --- | --- | --- |
| Age | 68 (59, 75) | 67 (58, 74) | 71 (63, 79) | <0.001 |
| Male | 19820 (59.2%) | 14832 (57.8%) | 4988 (64.0%) | <0.001 |
| Hypertension | 25982 (78.0%) | 19400 (75.8%) | 6582 (85.3%) | <0.001 |
| Diabetes | 11907 (35.6%) | 8451 (33.0%) | 3456 (44.4%) | <0.001 |
| Dyslipidemia | 24351 (73.1%) | 18571 (72.6%) | 5780 (74.9%) | <0.001 |
| Family History | 9510 (28.7%) | 7520 (29.6%) | 1990 (26.0%) | <0.001 |
| Smoking | 8802 (26.3%) | 6327 (24.7%) | 2475 (31.8%) | <0.001 |
| Previous CAD | 11184 (33.4%) | 7418 (28.9%) | 3766 (48.3%) | <0.001 |
| CAC score | 172 (0, 1051) | 110 (0, 837) | 523 (52, 1690) | <0.001 |
| Stress TPD | 4.28 (1.60, 10.4) | 3.60 (1.36, 8.51) | 7.59 (3.06, 17.7) | <0.001 |
| Rest MBF | 0.94 (0.74, 1.21) | 0.93 (0.73, 1.19) | 0.98 (0.74, 1.26) | <0.001 |
| Stress MBF | 2.19 (1.64, 2.80) | 2.30 (1.75, 2.88) | 1.82 (1.32, 2.46) | <0.001 |
| MFR | 2.30 (1.80, 2.87) | 2.42 (1.94, 2.98) | 1.88 (1.45, 2.40) | <0.001 |

**Table 2:** Characteristics of patients from the national lung cancer screening trial. CV – cardiovascular, COPD – chronic obstructive pulmonary disease, IQR – interquartile range

|  | **All Patients**<br>**N=24368** | **No CV Death**<br>**N=23909** | **CV Death**<br>**N=459** | **p-value** |
| --- | --- | --- | --- | --- |
| Age, median (IQR) | 61 (57, 65) | 60 (57, 65) | 64 (59, 68) | <0.001 |
| Male, n (%) | 14455 (59.3%) | 14122 (59.1%) | 333 (72.5%) | <0.001 |
| Pack years smoking, median (IQR) | 48 (39, 66.25) | 48 (39, 66) | 55 (44, 80) | <0.001 |
| Past Medical History, n (%) |  |  |  |  |
| Hypertension | 8552 (35.1%) | 8326 (34.8%) | 226 (49.2%) | <0.001 |
| Diabetes | 2362 (9.7%) | 2274 (9.5%) | 88 (19.2%) | <0.001 |
| Heart disease | 3161 (13.0%) | 3026 (12.7%) | 135 (29.4%) | <0.001 |
| COPD | 1248 (5.1%) | 1209 (5.1%) | 39 (8.5%) | <0.001 |
| Stroke | 681 (2.8%) | 642 (2.7%) | 39 (8.5%) | <0.001 |

### Chamber Density

LV chamber density was lower in patients with elevated resting MBF (defined as >1.2 mL/min/g), compared to patients with normal resting MBF (median 38 HU vs 40 HU, p<0.001). A full summary of chamber densities, and differences between myocardium and chamber density, stratified by elevated resting MBF is shown in **Supplemental Table 1**. The distribution of chamber densities and differences between chamber density and myocardium from the REFINE PET population are shown in **Supplemental Figure 1 and 2**, respectively. Two case examples are shown in **Figure 1**.

**Figure 1:**
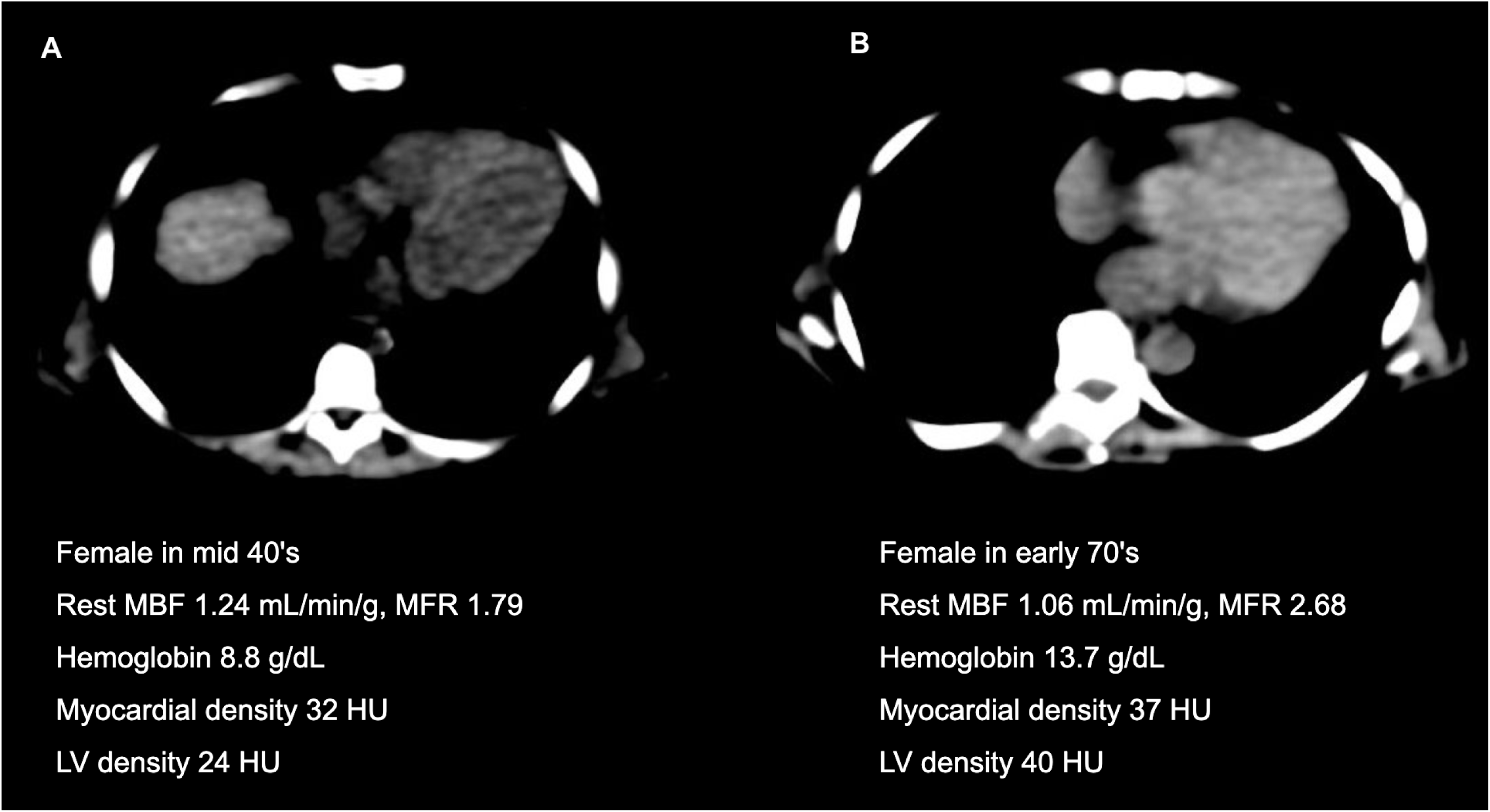
Case examples showing chamber density differences. Both computed tomography scans have a window level of 50 Hounsfield units (HU) and width of 70 HU. Images were denoised with a 2mm Gaussian filter. The patient in panel A had median myocardial density greater than median left ventricle (LV) density. The same patient had elevated resting myocardial blood flow (MBF), reduced myocardial flow reserve (MFR) and significant anemia. That patient experienced myocardial infarction (MI) 326 days after positron emission tomography (PET). The patient in panel B had median myocardial density lower than LV cavity, with normal MFR and no events during follow-up.

### Associations with Hemoglobin and Hematocrit

Hemoglobin and hematocrit values were available for 11353 patients. The median hemoglobin was 12.7 g/dL (IQR 11.2 – 14), with anemia present in 4955 (43.6%) and significant anemia (hemoglobin < 10 g/dL) in 1318 (11.6%). Patients who were classified as anemic had lower LV chamber density compared to patients who were not anemic (median 37 HU vs 40 HU, p<0.001). A similar difference was seen for other chamber densities as outlined in **Supplemental Table 2**.

Chamber density was positively correlated with hemoglobin (r 0.18 to 0.29, p<0.01 for all) and the difference between myocardium and chamber density was negatively correlated (r -0.19 to - 0.31, p<0.01 for all). All pairwise correlations as shown in **Figure 2**. The relationship between hemoglobin and difference between myocardium and LV density is shown in **Supplemental Figure 3** and diagnostic performance for significant anemia is shown in **Supplemental Table 3.**

**Figure 2:**
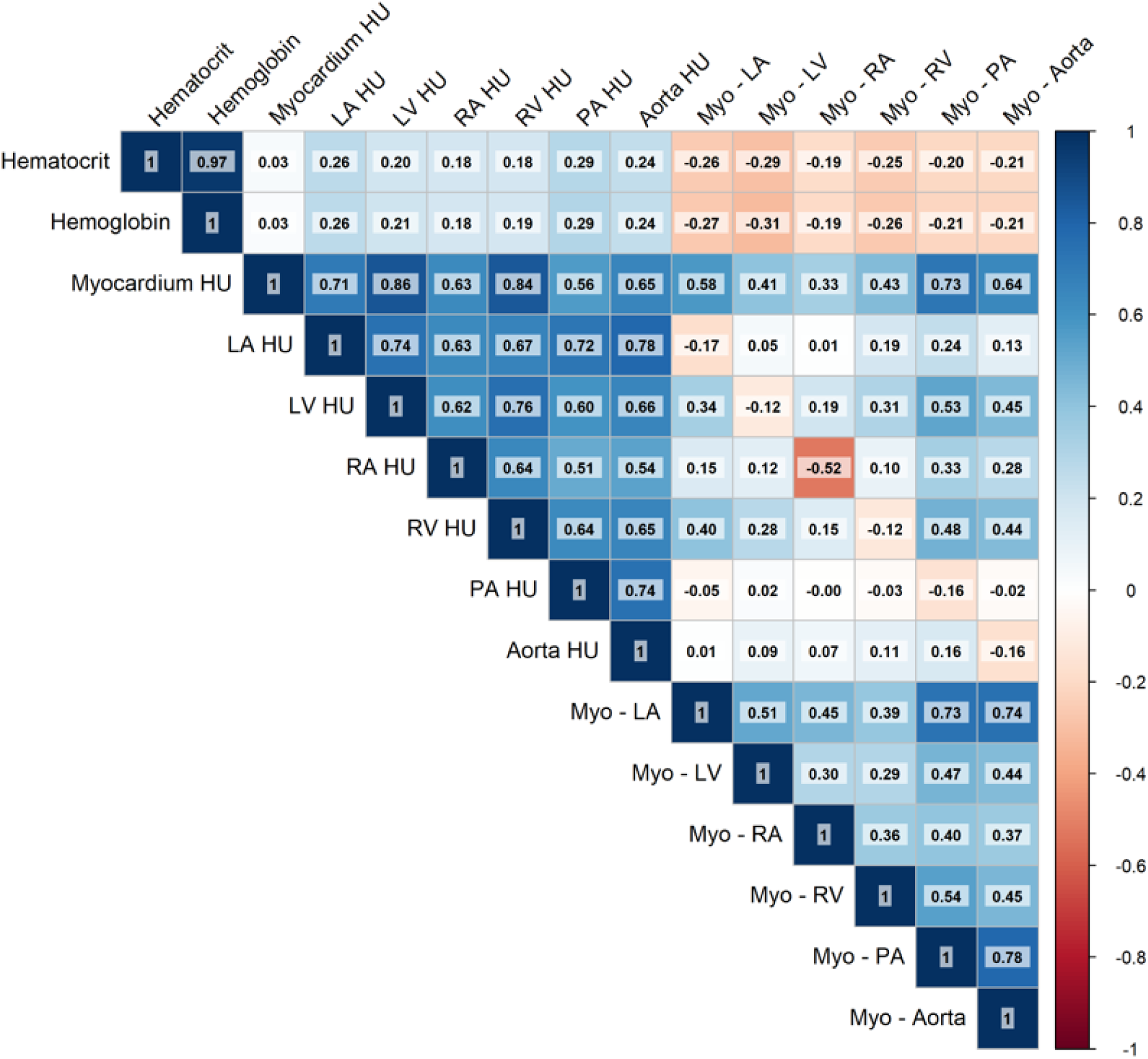
Correlation between blood pool density and difference between myocardium and blood pool density with hemoglobin and hematocrit in patients with available laboratory results. HU – Hounsfield Unit, LA – left atrium, LV – left ventricle, RA – right atrium, RV – right ventricle.

### Associations with Rest MBF and MFR

Higher LV density was associated with a lower likelihood of having high resting MBF (adjusted odds ratio [OR] per SD increase 0.41, 95% CI 0.32 – 0.52, p<0.001). The difference between myocardium and RV was also associated with the presence of high resting MBF (adjusted OR 1.45 per SD increase, 95% CI 1.31 – 1.61, p<0.001). Associations between high rest MBF and chamber density are shown in **Table 4**. Similarly, associations were identified with reduced MFR (**Supplemental Table 4)**. The association between myocardium to LV blood pool difference, modeled as a non-linear continuous variable, and reduced MFR, is shown in **Figure 3**. Correlations between chamber density measurements with rest MBF, and MFR are shown in **Supplemental Figure 4.**

**Figure 3.**
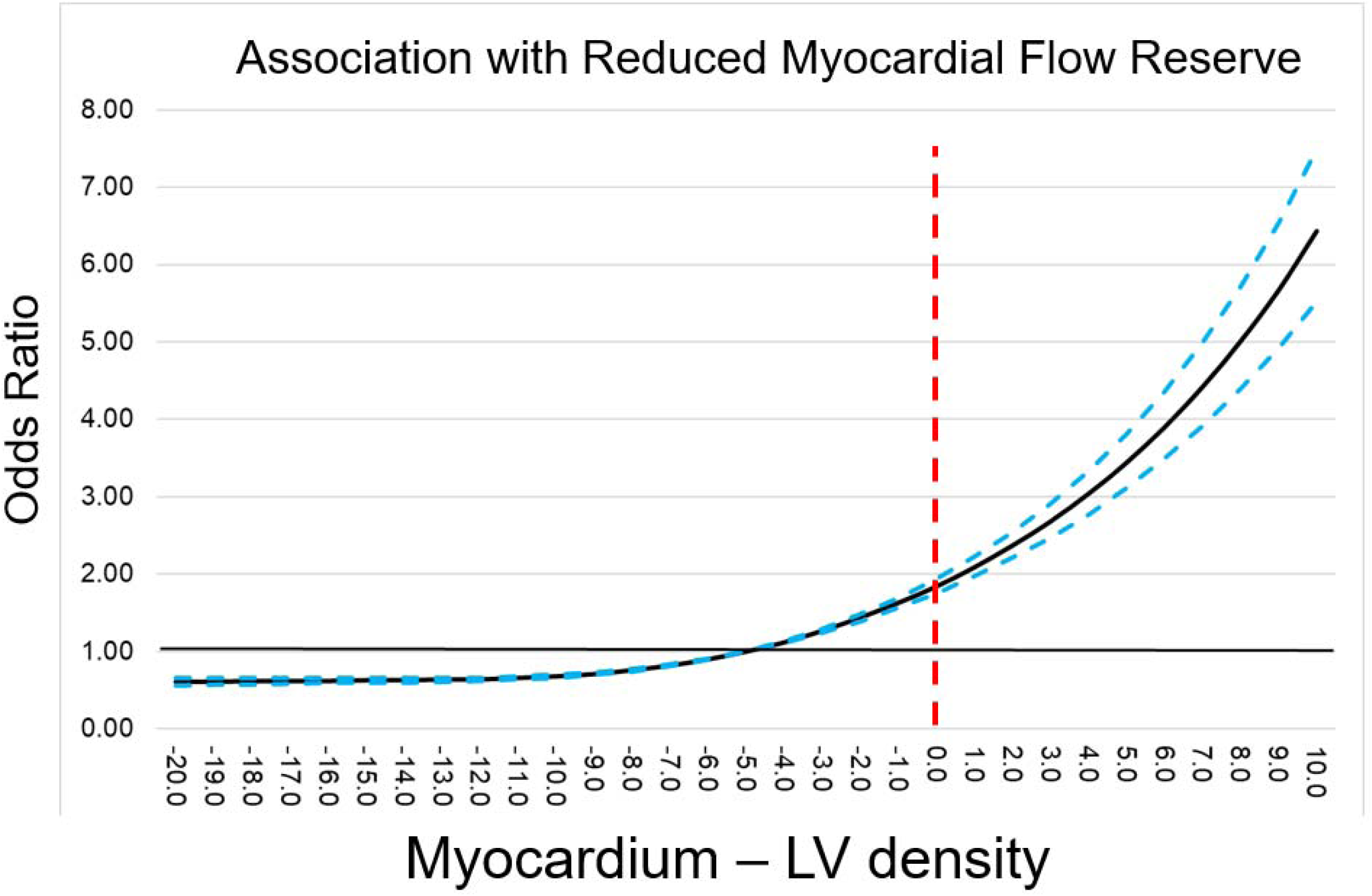
Relationship between the difference in median density of the myocardium and left ventricle (LV) and the likelihood of reduced myocardial flow reserve.

### Associations with Clinical Outcomes – REFINE PET

During median follow-up of 1511 days (IQR 769 – 2033), death or MI occurred in 7790 patients. Of those, 1783 patients experienced MI and 6582 patients died. Patients who experienced death or MI had lower median LV density (median 39 vs 40, p<0.001) and lower median LA HU (median 35 vs 37, p<0.001). A full summary of chamber density stratified by the incidence of death or MI is shown in **Table 3**.

**Table 3:**
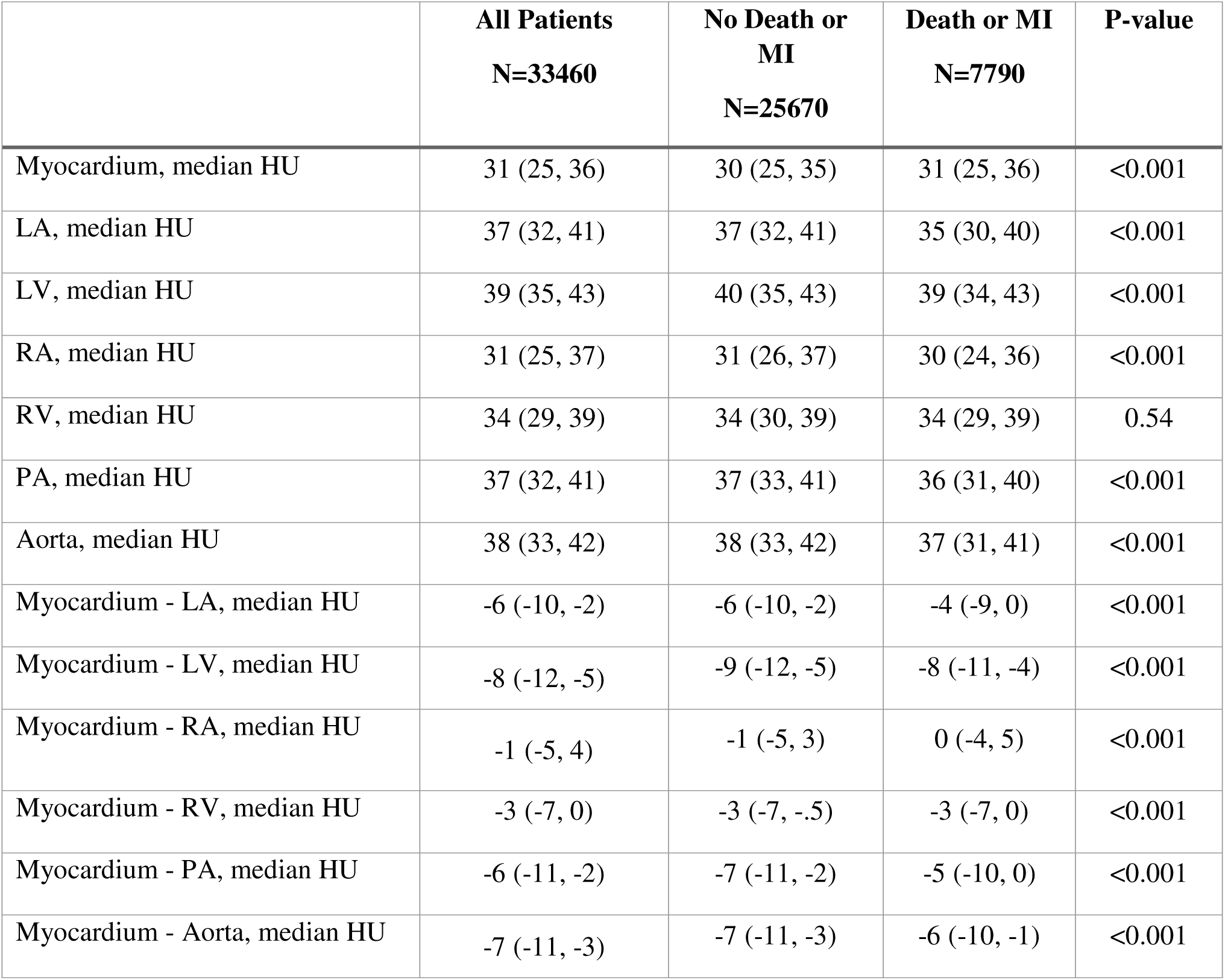
Chamber density in Hounsfield units (HU) stratified by incidence of death or myocardial infarction (MI)

**Table 4:**
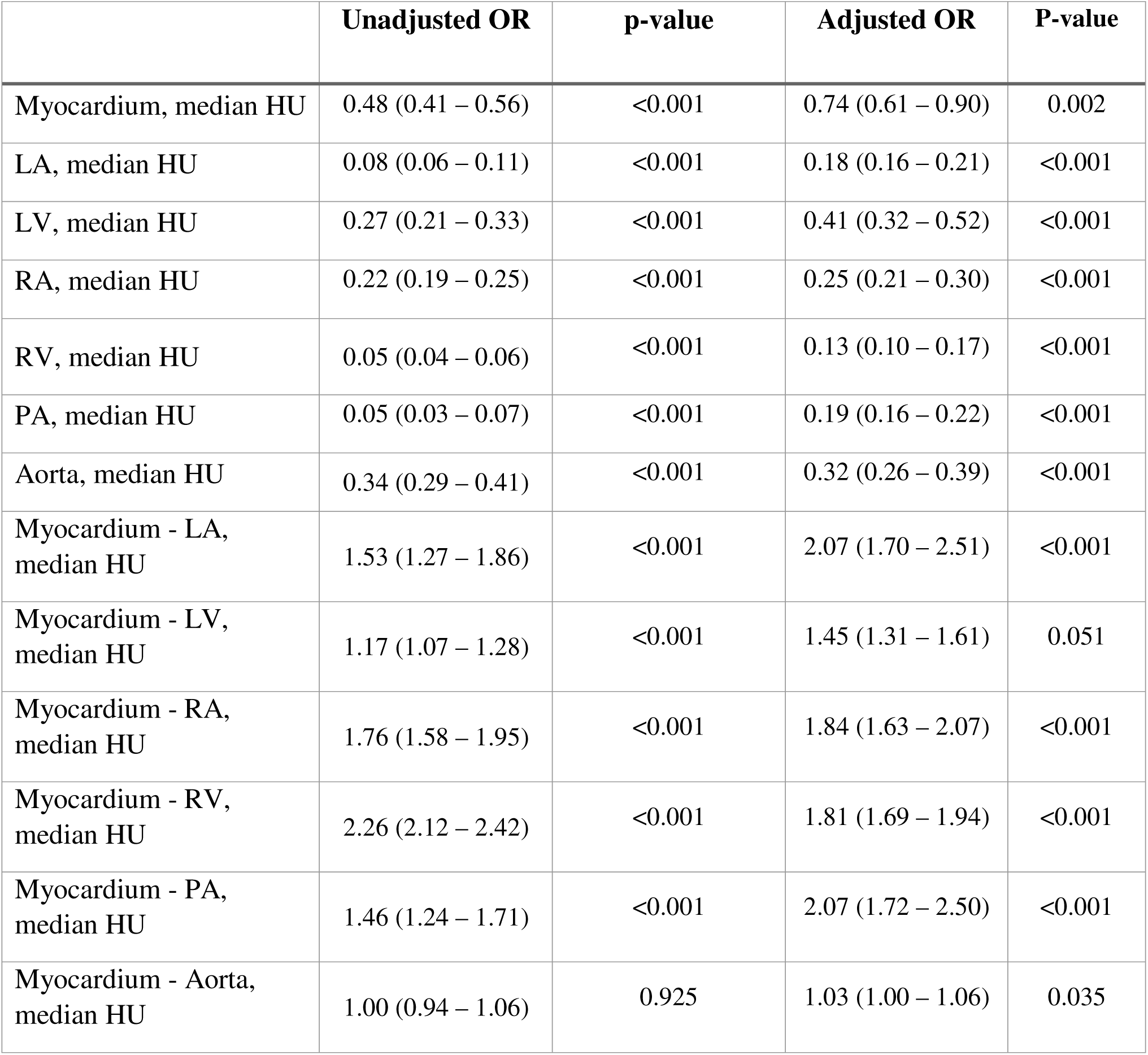
Associations between chamber densities and high resting myocardial blood flow (MBF). High resting MBF was defined as resting MBF > 1.2 mL/min/g. Multivariable model includes age, sex, body mass index, medical history, stress total perfusion deficit, coronary artery calcium, and left ventricular ejection fraction. To account for heterogeneity, site was incorporated as a random effect. Odds ratios reflect risk per standard deviation increase.

Associations between chamber density measures and the primary composite outcome are shown in **Table 5**. Higher LV density was associated with a decreased risk of death or MI in adjusted analyses (adjusted hazard ratio [HR] per SD increase 0.51, 95% CI 0.43 – 0.62). A higher difference between myocardium and LV density was also associated with increased risk (adjusted HR per SD increase 1.38, 95% CI 1.26 – 2.51). Incidence of death or MI, stratified by difference between myocardium and LV cavity as well as MFR, is shown in **Figure 4**. Patients with abnormal MFR and an increased difference between myocardium and LV blood pool were at the highest risk of death or MI (unadjusted HR 4.65, 95% CI 4.10 – 5.30, p<0.001). This difference persisted after adjusting for age, sex, past medical history, stress TPD, LVEF, CAC, and MFR (adjusted HR 2.54, 95% CI 2.21 – 2.91, p<0.001).

**Table 5:** Associations between chamber densities and incidence of death or myocardial infarction (MI). Multivariable model includes age, sex, body mass index, medical history, stress total perfusion deficit, myocardial flow reserve, coronary artery calcium, and left ventricular ejection fraction. Site was incorporated as a random effect using shared frailty models. Hazard ratios reflect risk per standard deviation increase.

|  | **Unadjusted HR** | **p-value** | **Adjusted HR** | **P-value** |
| --- | --- | --- | --- | --- |
| Myocardium median HU | 1.00 (0.92 – 1.02) | 0.836 | 0.79 (0.68 – 0.92) | 0.003 |
| LA median HU | 0.53 (0.43 – 0.65) | <0.001 | 0.35 (0.28 – 0.44) | <0.001 |
| LV median HU | 0.87 (0.73 – 1.02) | 0.094 | 0.51 (0.43 – 0.62) | <0.001 |
| RA median HU | 0.98 (0.94 – 1.03) | 0.433 | 0.76 (0.68 – 0.84) | <0.001 |
| RV median HU | 0.99 (0.95 – 1.02) | 0.513 | 0.56 (0.46 – 0.69) | <0.001 |
| PA median HU | 0.45 (0.35 – 0.57) | <0.001 | 0.41 (0.31 – 0.54) | <0.001 |
| Aorta median HU | 0.85 (0.74 – 0.98) | 0.026 | 0.59 (0.50 – 0.69) | <0.001 |
| Myocardium - LA median HU | 1.07 (1.04 – 1.09) | <0.001 | 1.05 (1.01 – 1.10) | 0.021 |
| Myocardium - LV median HU | 2.26 (2.09 – 2.45) | <0.001 | 1.38 (1.26 – 1.51) | <0.001 |
| Myocardium - RA median HU | 1.02 (1.01 – 1.04) | 0.009 | 1.01 (0.98 – 1.04) | 0.447 |
| Myocardium – RV median HU | 1.49 (1.40 – 1.59) | <0.001 | 1.11 (1.05 – 1.17) | <0.001 |
| Myocardium - PA median HU | 2.94 (2.55 – 3.40) | <0.001 | 1.20 (1.03 – 1.40) | <0.001 |
| Myocardium – Aorta median HU | 1.02 (1.00 – 1.03) | 0.010 | 1.02 (0.99 – 1.05) | 0.274 |

**Figure 4:**
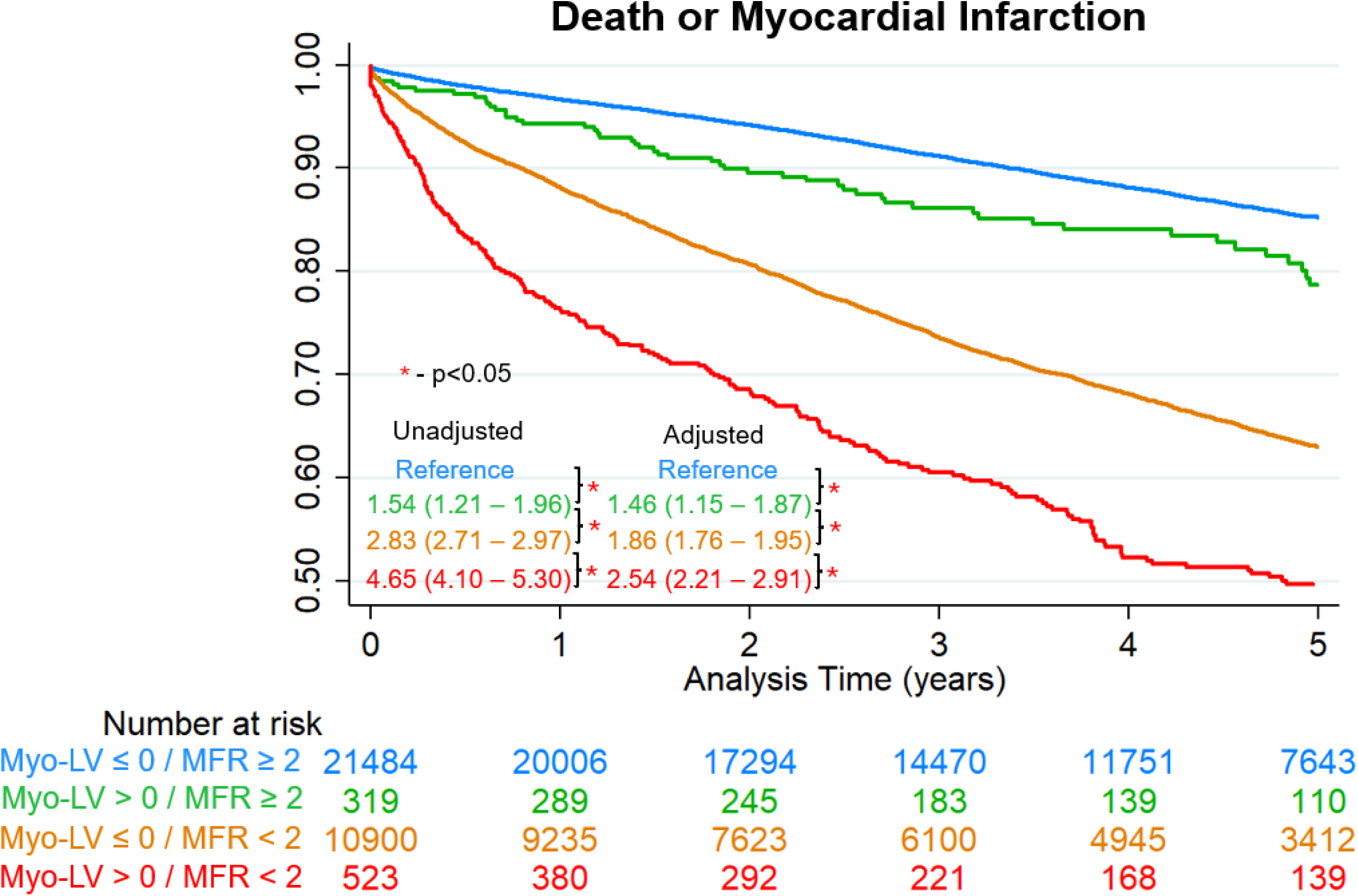
Associations with death or myocardial infarction (MI)in the positron emission tomography cohort. Difference was calculated between the myocardium and left ventricular blood pool, with differences > 0 considered significant. MFR – myocardial flow reserve.

### Associations with Outcomes in NLST Population

During a median follow-up of 6.7 (6.2 – 7.0) years, 459 (1.9%) patients experienced cardiovascular death. Patients with myocardium density greater than LV chamber density were at an increased risk of cardiovascular death (unadjusted HR 1.52, 95% CI 1.06 – 2.19, p=0.024) as shown in **Figure 5**. This risk persisted after adjustment for age, sex, smoking history and medical history (adjusted HR 1.71, 95% CI 1.18 – 2.47, p=0.004). All associations between chamber density measurements with cardiovascular mortality are outlined in **Supplemental Table 5.** Difference between myocardium and blood pool density was consistently associated with cardiovascular death, with adjusted HR ranging from 1.23 to 1.71.

**Figure 5:**
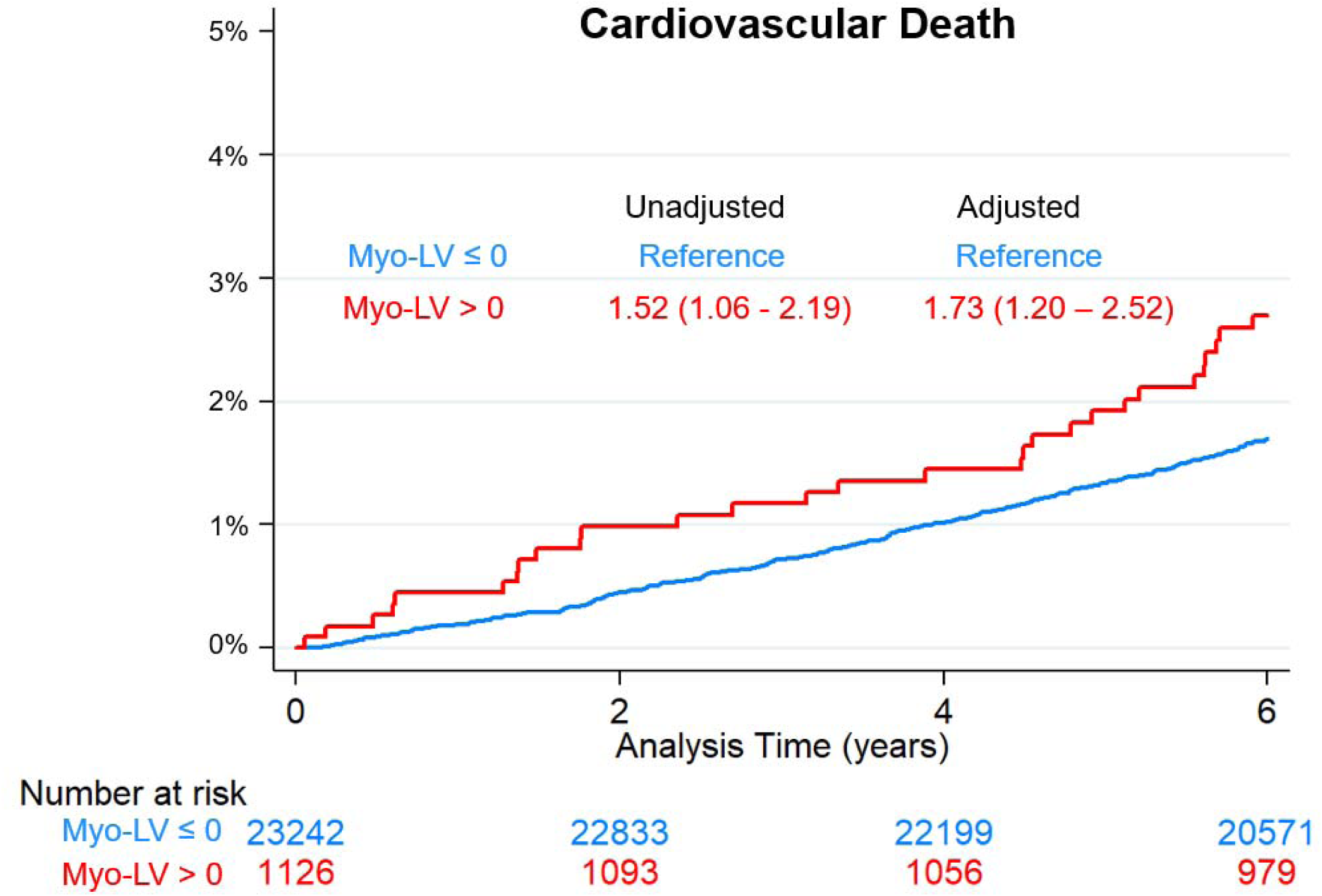
Associations with cardiovascular death in patients from the National Lung Cancer Screening trial (N =24368). Difference was calculated between the myocardium and left ventricular (LV) blood pool.

## DISCUSSION

We performed the largest analysis to date – including two large, multicenter datasets, reflecting a total of 57,828 patients - to evaluate the potential utility of cardiac chamber attenuation quantification. In patients undergoing PET MPI, we demonstrated that blood pool density was correlated with hemoglobin levels and associated with both resting myocardial blood flow (MBF) and myocardial flow reserve (MFR). More specifically, lower blood pool density and myocardial density higher than blood pool were associated with elevated resting MBF and reduced MFR. We went on to demonstrate that measures of blood pool density were associated with long-term cardiovascular outcomes after adjusting for age, sex, medical history, MFR, and CAC. Lastly, we demonstrated that this approach could be generalized to non-cardiac CT imaging. In patients undergoing lung cancer screening CT, a greater difference between myocardium and blood pool density was consistently associated with an increased risk of cardiovascular death. These findings provide large-scale, multicenter, and cross-modality validation of cardiac chamber density measurements as a method to extract physiologic information which can inform myocardial perfusion assessment and cardiovascular risk stratification.

In almost 11,353 patients with both hemoglobin measurements and CT images, we demonstrated a consistent relationship between blood pool density measurements with anemia. We identified consistent correlations to support the biological premise that reduced hemoglobin concentration leads to lower blood pool attenuation on non-contrast CT. This is consistent with previous studies evaluating chamber density (10–12). Abbasi et al. demonstrated coefficients of determination as high as 0.61. However, this was in a smaller population (n=325), with a wider distribution of hemoglobin (mean value 9.3 g/dL), manual measurements, and consistent imaging protocols(10). In contrast, we utilized automated measures in a much larger patient population which included a smaller proportion of anemic patients. While this likely led to lower correlation, it is more reflective of applying this approach in a general population. In spite of this, we also demonstrated good prediction performance for identifying patients with significant anemia. In fact, minimal differences between myocardium and blood pool density (which would not be readily apparent except with dedicated windowing) were highly specific (89-93%) for identifying patients with significant anemia. The use of DL-based segmentation enables automated, reproducible quantification across large populations, overcoming the limitations of subjective visual signs used previously.

In addition to correlations with laboratory measures of anemia, we evaluated the potential impact of these imaging markers of anemia on MBF. Patients with elevated resting MBF (>1.2 mL/min/g) had consistently lower blood pool density and higher myocardium density relative to blood pool. These provide a direct physiologic link between blood pool density as a surrogate of anemia and its impact on MBF which is consistent with previous studies evaluating impact of laboratory measures of anemia(9). We went on to demonstrate that chamber density measures were independently associated with high resting MBF and reduced MFR after adjusting for relevant confounders including age and medical history.

Lastly, we evaluated the potential utility of blood pool density measures for evaluating cardiovascular risk. We demonstrated that higher blood pool density was associated with lower risk of death or MI, while a larger myocardium–blood pool attenuation difference was associated with increased risk. Importantly, these attenuation biomarkers remained independently associated with adverse cardiovascular outcomes after adjustment for MFR, CAC, perfusion abnormalities, and clinical risk factors, suggesting that they capture biologically relevant cardiovascular risk not fully represented by existing imaging metrics. We also demonstrated and increased risk for cardiovascular death in patients with abnormal myocardium-blood pool differences in a second imaging modality. These two results in combination suggest that blood pool density measures could be an important new biomarker applied both to cardiac and non-cardiac imaging.

The potential utility of this approach extends to a large number of imaging tests. In PET MPI, identification of low chamber density could alert clinicians to the presence of anemia that might otherwise confound interpretation of MBF. However, CTAC scans are routinely acquired not only for PET but also for hybrid SPECT/CT imaging(29), where these measures could also be used to inform risk estimation. Since this method relies solely on non-contrast CT attenuation values (and automated chamber segmentation) it could be applied in all chest CT examinations with over 20 million performed annually in the United States(30). We demonstrated this generalizability by evaluating the utility of abnormal measurements (using thresholds derived in the PET MPI populations for a different outcome) in patients from the NLST. Importantly, anemia is a dynamic condition that can vary from mild to severe over time and opportunistic identification through CT could identify changes that are not apparent from prior laboratory imaging. Identification of patients with anemia could help physicians identify patients with subclinical cardiovascular disease or underlying inflammatory disease that require specific interventions or enhanced surveillance.

Several limitations should be acknowledged. Hemoglobin data were available only in a subset of patients and there is likely some selection bias inherent in which patients had available data. However, the median hemoglobin, and proportion of patients with significant anemia, in our analysis is likely more reflective of a typical referral population compared to prior analyses. CT acquisition parameters varied across sites, which may influence absolute attenuation values, although the large multicenter design enhances generalizability. As an observational study, causal relationships between chamber density, anemia, perfusion abnormalities, and outcomes cannot be definitively established. Finally, although attenuation differences were independently prognostic, formal assessment of incremental predictive value beyond established risk models warrants further investigation.

## CONCLUSION

In this large multicenter study of nearly 58,000 patients, including two imaging modalities, we demonstrate that CT images contain clinically important physiologic information related to anemia, MBF, and cardiovascular risk. These imaging biomarkers can be quantified opportunistically to provide additional physiologic and prognostic information from routinely acquired CT data to help guide clinical care.

## Data Availability

To the extent allowed by data and code sharing agreements and IRB protocols, the deidentified data and analysis code from this manuscript will be shared upon written request.

## CLINICAL PERSPECTIVES

### Competency in Medical Knowledge

Cardiac chamber densities on computed tomography are correlated with hemoglobin and myocardial blood flow and are associated with cardiovascular risk.

### Translational Outlook

Prospective studies evaluating whether these measures can be used to improve interpretation of positron emission tomography or guide medical therapy are warranted.

## ABBREVIATIONS

AI: artificial intelligence
CAD: coronary artery disease
CT: computed tomography
CTAC: computed tomography attenuation correction
DL: deep learning
MFR: myocardial flow reserve
MPI: myocardial perfusion imaging
PET: positron emission tomography
RV: right ventricle
SPECT: single photon emission computed tomography

## Central Illustration

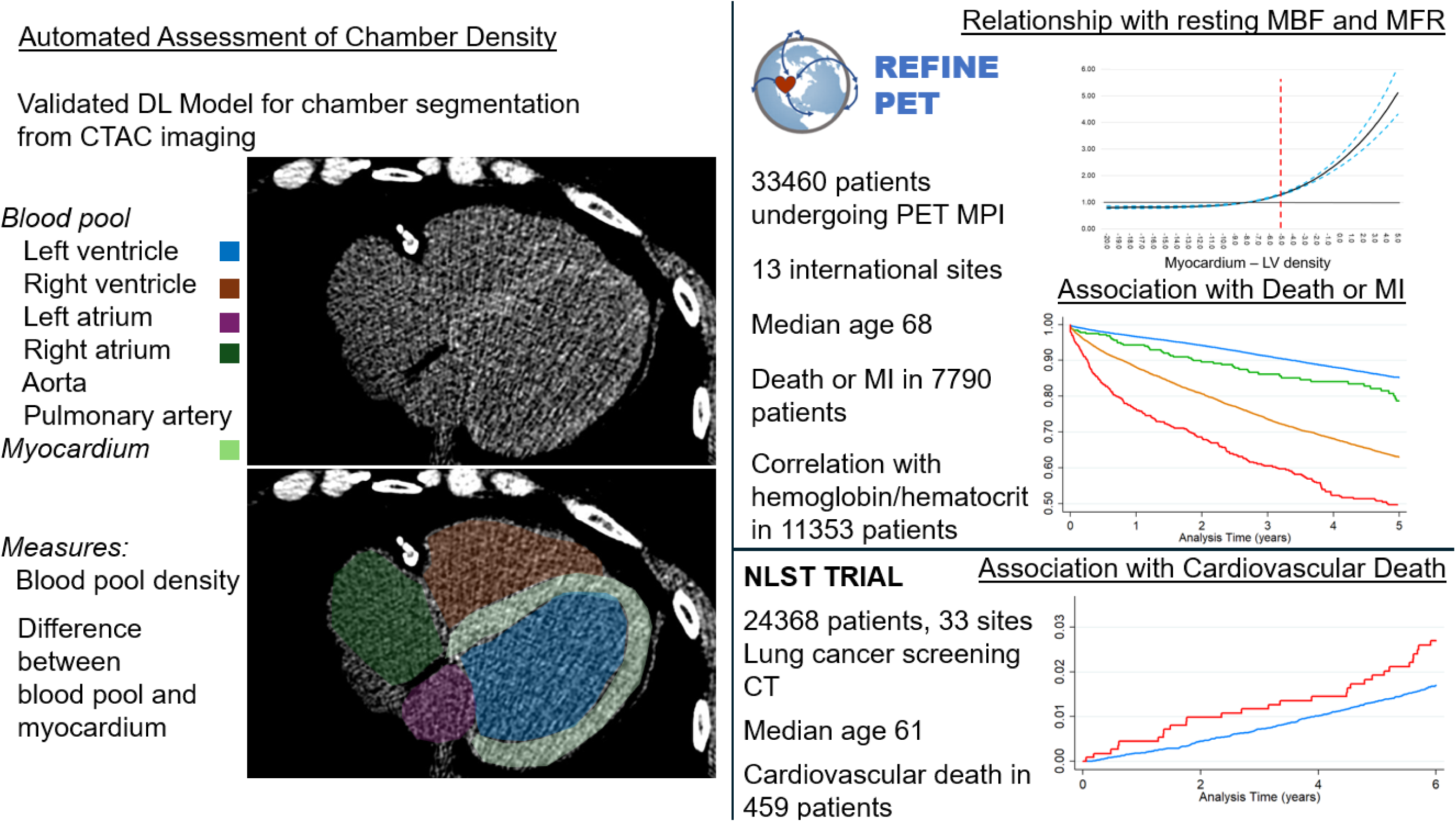

