## Supplemental Material for "Opportunistic CT Attenuation Biomarkers of Anemia Are Associated With Impaired Myocardial Flow Reserve and Cardiovascular Outcomes"

**SUPPLEMENT**

|  | All Patients  N=33460 | Normal resting MBF (<1.2)  N=24770 | High resting MBF (>1.2)  N=8690 | P-value |
| --- | --- | --- | --- | --- |
| Myocardium, median HU | 31 (25, 36) | 31 (26, 36) | 30 (24, 35) | <0.001 |
| LA, median HU | 37 (32, 41) | 38 (32, 42) | 35 (30, 40) | <0.001 |
| LV, median HU | 39 (35, 43) | 40 (35, 43) | 38 (34, 42) | <0.001 |
| RA, median HU | 31 (25, 37) | 32 (26, 37) | 30 (23, 35) | <0.001 |
| RV, median HU | 34 (29, 39) | 35 (30, 40) | 32 (27, 37) | <0.001 |
| PA, median HU | 37 (32, 41) | 37.5 (33, 41) | 36 (31, 40) | <0.001 |
| Aorta, median HU | 38 (33, 42) | 38 (33, 42) | 37 (32, 41) | <0.001 |
| Myocardium - LA, median HU | -6 (-10, -2) | -6 (-10, -2) | -6 (-10, -1) | <0.001 |
| Myocardium - LV, median HU | -8 (-12, -5) | -8 (-12, -5) | -9 (-12, -5) | 0.001 |
| Myocardium - RA, median HU | -1 (-5, 4) | -1 (-5, 3) | 0 (-4, 5) | <0.001 |
| Myocardium - RV, median HU | -3 (-7, 0) | -4 (-7, -1) | -2 (-6, 1) | <0.001 |
| Myocardium - PA, median HU | -6 (-11, -2) | -6 (-11, -2) | -6 (-11, -1) | 0.62 |
| Myocardium - Aorta, median HU | -7 (-11, -3) | -7 (-11, -3) | -7 (-12, -3) | <0.001 |

Supplemental Table 1: Chamber density stratified by the presence of high resting myocardial blood flow (MBF). High resting MBF was defined as resting MBF > 1.2 mL/min/g.

|  | All Patients  N=11353 | Not Anemic  N=6398 | Anemic  N=4955 | P-value |
| --- | --- | --- | --- | --- |
| Myocardium, median HU | 30 (24, 35) | 30 (24, 35) | 30 (24, 35) | 0.32 |
| LA, median HU | 35 (30, 40) | 37 (32, 41) | 33 (28, 38) | <0.001 |
| LV, median HU | 39 (34, 43) | 40 (35, 44) | 37 (32, 42) | <0.001 |
| RA, median HU | 29 (23, 35) | 31 (24.5, 36) | 28 (22, 33) | <0.001 |
| RV, median HU | 34 (28, 38) | 35 (30, 39) | 32 (27, 37) | <0.001 |
| PA, median HU | 35 (30, 39) | 36 (32, 40) | 33 (28, 37) | <0.001 |
| Aorta, median HU | 36 (31, 41) | 38 (33, 42) | 34 (29, 39) | <0.001 |
| Myocardium - LA, median HU | -5 (-10, -1) | -7 (-11, -3) | -3 (-7, 1) | <0.001 |
| Myocardium - LV, median HU | -9 (-12, -6) | -10 (-13, -7) | -7 (-11, -4) | <0.001 |
| Myocardium - RA, median HU | 1 (-4, 6) | 0 (-5, 4) | 2 (-3, 7) | <0.001 |
| Myocardium - RV, median HU | -4 (-7, -1) | -5 (-8, -2) | -3 (-6, 1) | <0.001 |
| Myocardium - PA, median HU | -5 (-10, 0) | -6 (-11, -2) | -3 (-8, 2) | <0.001 |
| Myocardium - Aorta, median HU | -6 (-11, -2) | -7 (-12, -3) | -4 (-9, 0) | <0.001 |

Supplemental Table 2: Chamber density measurements in patients with available hemoglobin data. Patients were classified as having anemia based on hemoglobin <13.2 g/dL in male patients or <11.6 g/dL in female patients.

|  | Threshold | Sensitivity | Specificity |
| --- | --- | --- | --- |
| LA, median HU | 35 | 69 | 57 |
| LV, median HU | 35 | 45 | 74 |
| RA, median HU | 35 | 89 | 28 |
| RV, median HU | 35 | 72 | 48 |
| PA, median HU | 35 | 74 | 54 |
| Aorta, median HU | 35 | 60 | 63 |
| Myocardium - LA, median HU | 0 | 25 | 91 |
| Myocardium - LV, median HU | 0 | 47 | 89 |
| Myocardium - RA, median HU | 0 | 16 | 93 |
| Myocardium - RV, median HU | 0 | 26 | 92 |
| Myocardium - PA, median HU | 0 | 26 | 92 |
| Myocardium - Aorta, median HU | 0 | 22 | 91 |

Supplemental Table 3: Thresholds for chamber density measurements for predicting anemia.

|  | Unadjusted OR | p-value | Adjusted OR | P-value |
| --- | --- | --- | --- | --- |
| Myocardium, median HU | 1.00 (0.97 – 1.02) | 0.737 | 0.76 (0.64 – 0.91) | 0.003 |
| LA, median HU | 0.08 (0.07 – 0.11) | <0.001 | 0.15 (0.14 – 0.16) | <0.001 |
| LV, median HU | 0.28 (0.23 – 0.35) | <0.001 | 0.26 (0.23 – 0.31) | <0.001 |
| RA, median HU | 0.38 (0.33 – 0.43) | <0.001 | 0.22 (0.19 – 0.26) | <0.001 |
| RV, median HU | 0.35 (0.29 – 0.43) | <0.001 | 0.23 (0.20 – 0.36) | <0.001 |
| PA, median HU | 0.05 (0.03 – 0.06) | <0.001 | 0.13 (0.12 – 0.14) | <0.001 |
| Aorta, median HU | 0.30 (0.26 – 0.36) | <0.001 | 0.28 (0.24 – 0.31) | <0.001 |
| Myocardium - LA, median HU | 5.20 (4.40 – 6.14) | <0.001 | 2.86 (2.37 – 3.44) | <0.001 |
| Myocardium - LV, median HU | 3.50 (3.22 – 3.82) | <0.001 | 3.02 (2.72 – 3.38) | <0.001 |
| Myocardium - RA, median HU | 3.17 (2.87 – 3.50) | <0.001 | 2.20 (1.97 – 2.46) | <0.001 |
| Myocardium - RV, median HU | 1.96 (1.85 – 2.07) | <0.001 | 1.83 (1.72 – 1.95) | <0.001 |
| Myocardium - PA, median HU | 5.30 (4.56 – 6.17) | 0.673 | 3.93 (3.30 – 4.69) | <0.001 |
| Myocardium - Aorta, median HU | 3.43 (2.91 – 4.05) | <0.001 | 1.02 (0.99 – 1.05) | 0.136 |

Supplemental Table 4. Associations with reduced myocardial flow reserve, defined as values < 2. Multivariable model includes age, sex, body mass index, medical history, stress total perfusion deficit, coronary artery calcium, and left ventricular ejection fraction. Odds ratios reflect risk per standard deviation increase.

|  | Unadjusted HR | p-value | Adjusted HR | P-value |
| --- | --- | --- | --- | --- |
| LA median HU | 1.10 (0.88 – 1.38) | 0.421 | 0.96 (0.77 – 1.21) | 0.754 |
| LV median HU | 1.58 (0.51 – 4.91) | 0.431 | 1.32 (0.42 – 4.12) | 0.629 |
| RA median HU | 1.06 (0.87 – 1.29) | 0.552 | 1.00 (0.83 – 1.22) | 0.964 |
| RV median HU | 1.40 (1.15 – 1.71) | 0.001 | 1.30 (1.06 – 1.59) | 0.012 |
| PA median HU | 0.78 (0.53 – 1.16) | 0.219 | 0.88 (0.59 – 1.30) | 0.511 |
| Aorta median HU | 0.91 (0.60 – 1.38) | 0.652 | 1.03 (0.67 – 1.57) | 0.896 |
| Myocardium - LA median HU | 1.25 (1.02 – 1.52) | 0.028 | 1.23 (1.01 – 1.51) | 0.039 |
| Myocardium - LV median HU | 1.52 (1.06 – 2.19) | 0.024 | 1.71 (1.18 – 2.47) | 0.004 |
| Myocardium - RA median HU | 1.52 (1.26 – 1.84) | <0.001 | 1.46 (1.20 – 1.77) | <0.001 |
| Myocardium – RV median HU | 1.60 (1.31 – 1.95) | <0.001 | 1.68 (1.37 – 2.07) | <0.001 |
| Myocardium - PA median HU | 1.30 (1.00 – 1.68) | 0.050 | 1.47 (1.13 – 1.92) | 0.004 |
| Myocardium – Aorta median HU | 1.29 (1.01 – 1.66) | 0.042 | 1.48 (1.15 – 1.91) | 0.002 |

Supplemental Table 5. Associations with cardiovascular death in the national lung cancer screening trial. Multivariable model includes age, sex, smoking history and past medical history.

Supplemental Figure 1:

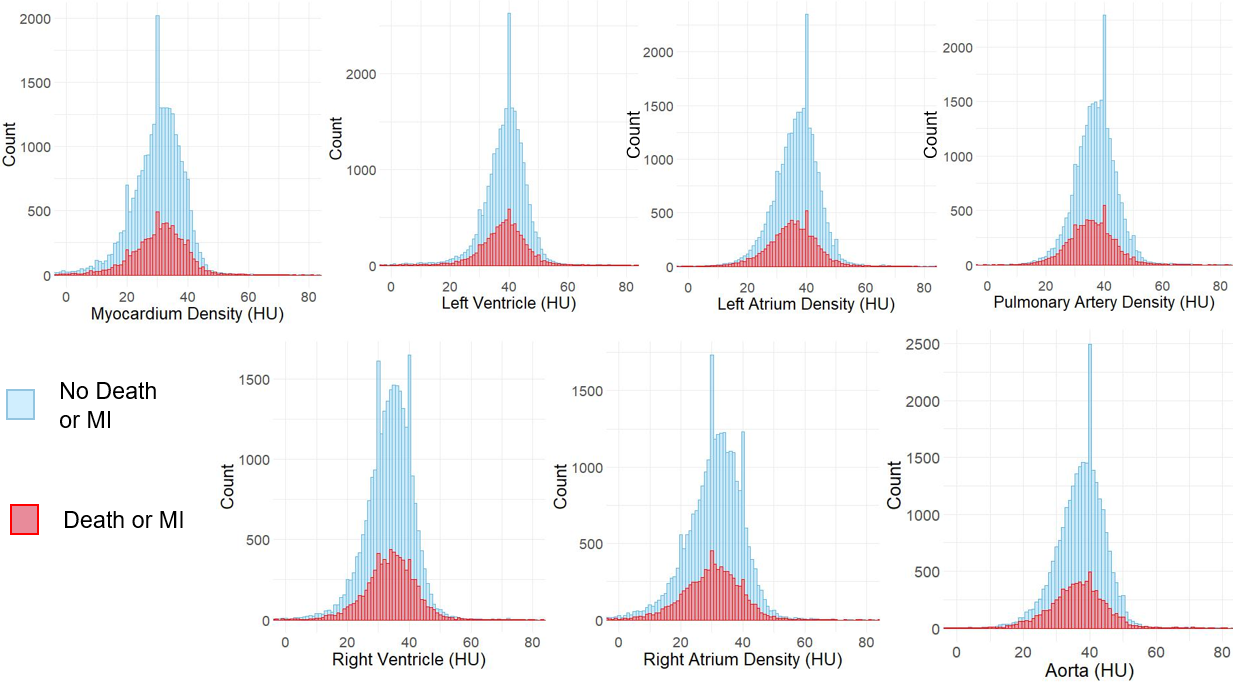

Supplemental Figure 1: Distribution of chamber density measurements. All densities are in Hounsfield units (HU). MI – myocardial infarction.

Supplemental Figure 2:

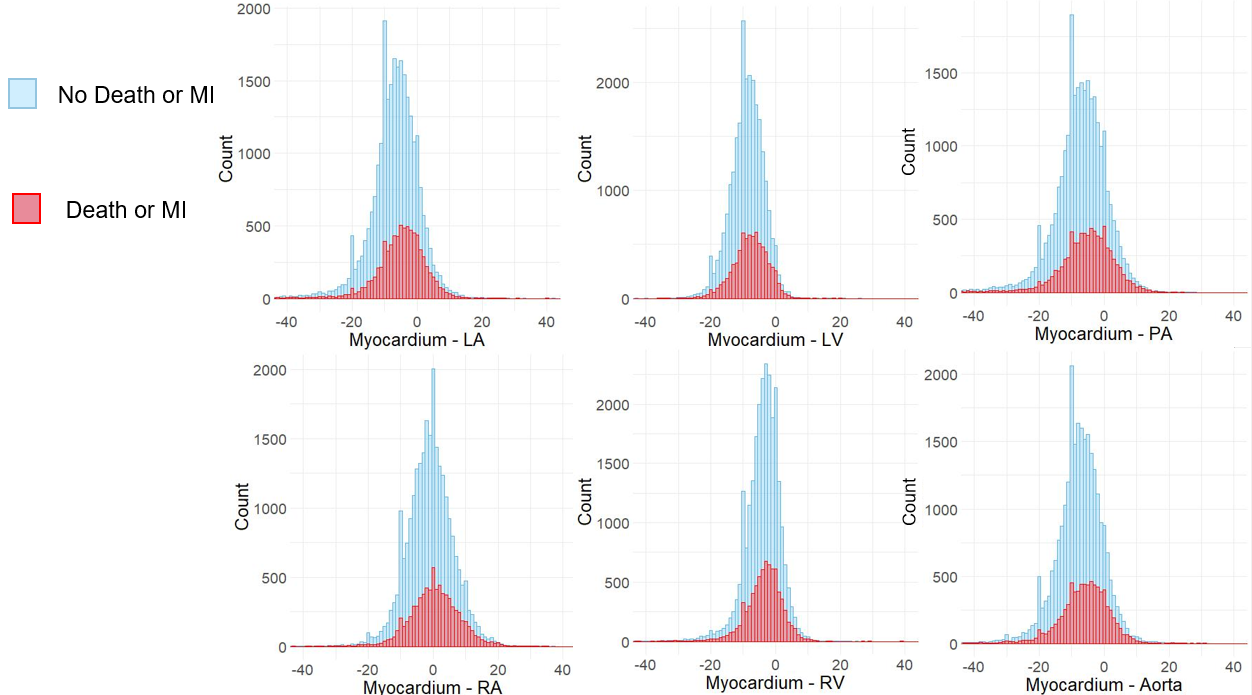

Supplemental Figure 2: Distribution of chamber density differences compared to myocardium. All densities are in Hounsfield units (HU). MI – myocardial infarction.

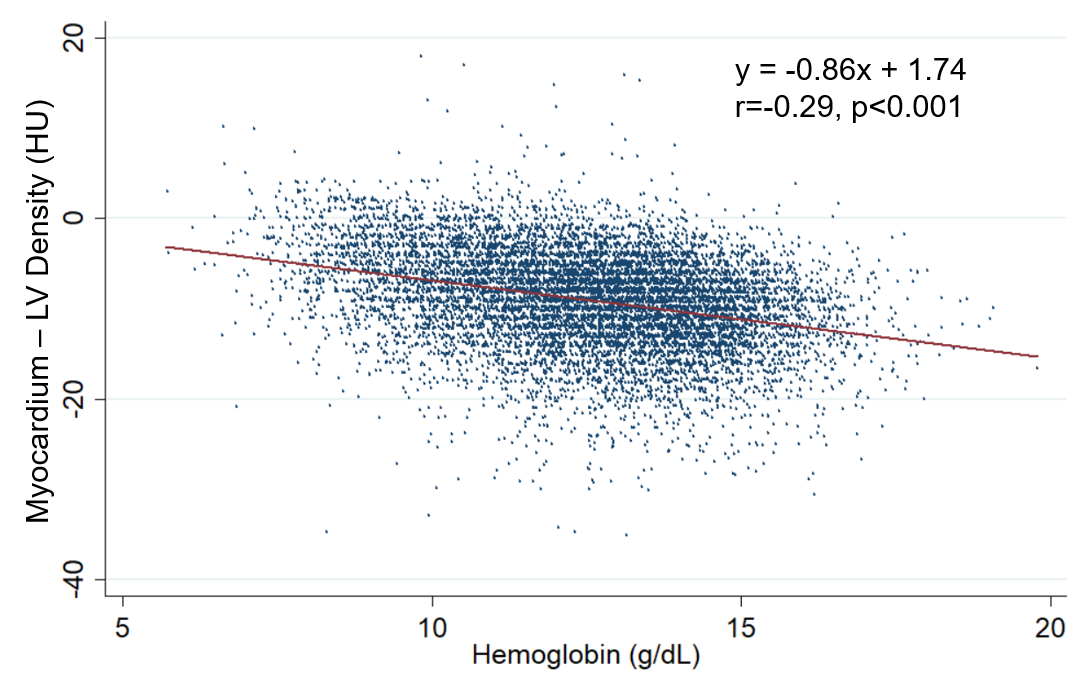

Supplemental Figure 3: Relationship between hemoglobin and the difference between myocardium and left ventricle (LV) cavity density. Analysis performed in 11353 patients with available laboratory data.

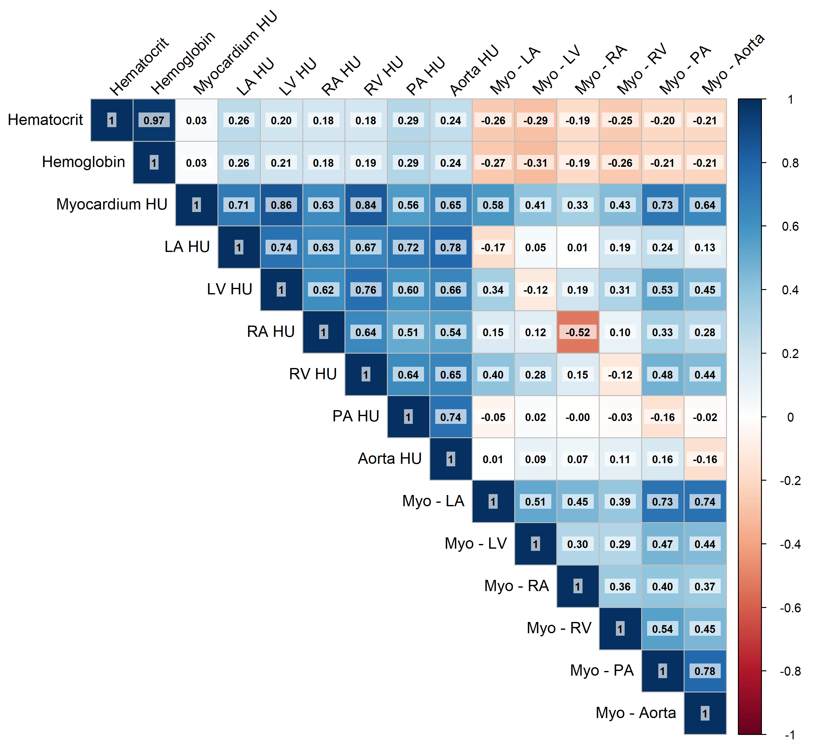

Supplemental Figure 4: Correlation between blood pool density and difference between myocardium and blood pool density with myocardial blood flow, and myocardial flow reserve.
